# Towards personalised early prediction of Intra-Operative Hypotension following anesthesia using Deep Learning and phenotypic heterogeneity

**DOI:** 10.1101/2023.01.20.23284432

**Authors:** Anna Tselioudis Garmendia, Ioannis Gkouzionis, Charalampos P. Triantafyllidis, Vasileios Dimakopoulos, Sotirios Liliopoulos, Dragana Vuckovic, Lucas Paseiro-Garcia, Marc Chadeau-Hyam

## Abstract

Intra-Operative Hypotension (IOH) is a haemodynamic abnormality that is commonly observed in operating theatres following general anesthesia and associates with life-threatening post-operative complications. Using Long Short Term Memory (LSTM) models applied to Electronic Health Records (EHR) and time-series intra-operative data in 604 patients that underwent colorectal surgery we predicted the instant risk of IOH events within the next five minutes. K-means clustering was used to group patients based on pre-clinical data. As part of a sensitivity analysis, the model was also trained on patients clustered according to Mean artelial Blood Pressure (MBP) time-series trends at the start of the operation using K-means with Dynamic Time Warping. The baseline LSTM model trained on all patients yielded a test set Area Under the Curve (AUC) value of 0.83. In contrast, training the model on smaller sized clusters (grouped by EHR) improved the AUC value (0.85). Similarly, the AUC was increased by 4.8% (0.87) when training the model on clusters grouped by MBP. The encouraging results of the baseline model demonstrate the applicability of the approach in a clinical setting. Furthermore, the increased predictive performance of the model after being trained using a clustering approach first, paves the way for a more personalised patient stratification approach to IOH prediction using clinical data.

## Introduction

Arterial hypotension is a highly prevalent haemodynamic abnormality that is commonly observed in patients undergoing surgery under general anesthesia (1, 2), and notably in colorectal surgery. The risk of resulting intra-operative hypotension (IOH), defined as Mean Blood Pressure (MBP) *<*65 mmHg (3), has been associated with many factors including age, comorbidity, type of anesthesia, and low systolic blood pressure (SBP) (4, 5). Anesthesia itself is thought to play a role in the occurrence of IOH. Anaesthetic agents, such as propofol, inhibit the sympathetic nervous system and impair the baroreflex regulatory mechanisms, both of which play a central role in blood pressure regulation(6). If not promptly treated, the decline of MBP can have adverse consequences as this compromises local tissue perfusion and can cause hypoxic damages in vital organs (7). IOH is also thought to promote local infection; a complication that is mostly observed during colorectal surgeries (8). Additionally, an increasing number of studies reveal a link between IOH and other life-threatening complications, such as myocardial infraction and acute kidney injury (9, 10, 11, 12, 13, 14, 15). This ultimately translates into increased in-hospital length of stay (LoS) and postoperative mortality and subsequent morbidity (16, 1).

Due to the complex pathophysiology underlying IOH, real-time prediction of the short term risk of the occurrence of such events while the patient is under anesthesia is deemed critical (17, 18). In the setting of a busy operating room, clinicians often have to manage the onset of IOH without additional warning. While standard patient monitoring devices measure instantaneous physiological signals including blood pressure, they do not account for temporal trends in vitals that may indicate earlier in time an increased and developing risk of the occurrence of an imminent IOH event (19). However, even a five minute time window warning of an imminent IOH event could lessen hypotension-associated postoperative complications (20), as it would allow anaesthesiologists sufficient time to proactively maintain the haemodynamic stability of the patient (e.g. by adjusting anesthetic agents, fluids and vasoactive drugs). Modern patient monitoring systems have been expanded to include data collected from different devices. Combined with the high-resolution physiological/pharmacological time-series data and Electronic Health Records (EHR) of the patient, more granular data could be integrated to better predict the occurrence of imminent IOH incidents in real-time (21).

In order to accommodate the size and the complexity of such medical data, the use of modern Machine Learning (ML) techniques appears as an important tool to support clinicians in the operating theater and guide their decisions in real-time. Supervised Machine Learning methods provide an opportunity to exploit large amounts of sequential data and are therefore increasingly utilized in training and analysing physiological timeseries for prediction. Deep learning (DL) methods also represent an important alternative to assist clinicians as they can learn more complex relationships of the input data without the need of human interpretation (22, 23, 20, 24, 25). In the field of anaesthesiology, the development of algorithms predictive of intra-operative physiological alterations is showing encouraging results, often outperforming traditional modelling (26, 27, 28).

Here, we propose a Long Short-Term Memory (LSTM, (29)) model to accurately predict the occurrence of imminent IOH events 5 minutes ahead and provide a time-resolved early warning risk score. To achieve this, the analysis uses EHR and physiological time-series data collected from 604 patients undergoing colorectal surgery under general anesthesia from the Vital Signs DataBase (VitalDB), published by Seoul National University Hospital (30). We additionally investigate whether the predictive performance of the baseline model (the model trained on all data) can be improved by training the model on different clusters of patients that have been partitioned according to their clinical characteristics and intra-operative vitals. More specifically, splitting the initial cohort of patients using clustering methods based on their clinical features and intra-operative vitals yields smaller clusters of the cohort that may be more suitable to be used independently in the training of ML models on them. For instance, increasing evidence suggests that factors such as male sex, increased BMI and age, as well as the American Society of Anaesthesiologists (ASA) Physical Status score IV (subjective assessment of a patient’s health before surgery) are linked to the occurrence of hypotensive events during colorectal surgery (1). Ultimately, the findings of this study aid in the development of a modelling framework for real-time anesthesia monitoring and prediction of IOH in a clinical setting, to proactively avert associated post-operative complications and mortality.

Patient physiological time-series data, defined as a collection of data points recorded at constant time intervals, represent an invaluable source of information that is under-exploited. Most of the studies for IOH prediction make use of such data by employing forecasting methods such as the Autoregressive Integrated Moving Average (ARIMA), which regresses the observed value of the variables on the previous values observed for that variable, and applies a weight to lagging observations in order to forecast future time-series values according to their inherent temporal nature and error terms. The word *integrated* refers to the removal of the stationary nature that usually characterises time-series data, while *moving average* here indicates the lag of the forecast errors in the moving average model. However, this method performs poorly with long term forecasting, as it assumes a linear relationship between independent and dependent variables and only works with univariate analysis. These limitations render ARIMA unsuitable for real-world problems, characterised by complex non-linear mechanisms (31). Additionally, a recent study (24) developed a hybrid Convolutional Neural Network (CNN) model, combining parameters extracted from invasive (arterial pressure waveform as a single input signal, and a multichannel model including arterial pressure, electrocardiography, photoplethysmography and capnography) and non-invasive patient monitoring, (photoplethysmography waveform as single input signal, and a multichannel model including other signals besides arterial pressure). It was utilised for the real-time prediction of a IOH at 5, 10 and 15 minutes in advance using combined biosignal waveforms (acquired using one-point SBP/DBP, photoplethysmography, capnography and electrocardiography) and may provide a more reliable prediction for patients than the more invasive arterial catheterisation. Another study (22) aimed at predicting hypotensive events occurring between tracheal intubation and incision. To do so, the authors trained several machine learning models with data recorded from the start of anesthesia induction to right before intubation. By adopting a feature engineering approach, a Random Forest (RF) approach combined with feature selection showed the highest accuracy, followed by CNN and XGBoost. Similarly, in (4) a predictive model was developed for anesthesia-induced hypotensive events with the same time constraints. Again, RF outperformed all other models, including Naïve Bayes, RNNs and logistic regression. However, both studies focus on patients having undergone laparoscopic cholecystectomy, while neither account for intra-operative hypotension. In contrast, (1) hypothesised that different phases of hypotension during an operation have distinct causative mechanisms. Therefore, they distinguished hypotension at two phases: post-induction hypotension (PIH) and early intra-operative hypotension (eIOH). Multivariate logistic regression was subsequently used to find clinical factors independently related to PIH and eIOH, supporting the hypothesis of there being a temporal component to the causative mechanisms of hypotension. Furthermore, (32) attempted multiple ML algorithms using features extracted from EHR, with the aim of predicting PIH within 10 minutes of anesthesia induction. The best performing model in post-induction hypotension prediction was tuned gradient boosting (AUC=0.76), followed by RF (AUC=0.74). Although the definition of hypotension was stricter than in most studies (MAP *<*55 mmHg), a sensitivity analysis using the conventional threshold at 65mmHg showed an AUC reduced to 0.72 for the best performing model. Notably, the authors used EHR data, as opposed to intra-operative vital signs data (32).

In (20), a weighted average ensemble of individual neural networks was used to predict five-minute IOH arterial waveforms (as opposed to our non-invasive MBP recording variable used to train the models), which performed worse than our suggested approach (precision-recall curve (AUPRC) score at best 0.72). Additionally, the *Hypotension Prediction Index* (HPI) (33, 34, 35), an algorithm for the real-time prediction of hypotension, constitutes the first clinically available application of IOH prediction. The index uses ML models to continuously analyse multiple haemodynamic features, yielding a unit-less number ranging from 0-100 that indicates the likelihood of an imminent hypotensive event. Following model validation, the algorithm predicted hypotension with a sensitivity and specificity of 89% and 90% respectively, 10 minutes before the onset of hypotension (19). In order to increase precision, the authors opted for a binary classification algorithm, during the development of which periods of hypotension were arbitrarily defined as MBP *<*65 mmHg (*>*1 minute), whereas periods of non-hypotension were defined by MBP *>*75 mmHg, creating a *gray zone* for MBP (ranging from 65 to 75 mmHg). Furthermore, the model did not investigate hypotensive events occurring during anesthesia induction. Lastly, although the algorithm was trained on mixed ICU/operating room patient data, it was tested on surgical patient data (36). On a higher level, existing literature uses arterial pressure vital signals as a single data source, whereas IOH is also linked to other physiological alterations, such as respiratory patterns and ECG trends (24).

The motivation behind LSTMs, introduced by (29), was to mitigate the difficulty of RNNs to process long-tern dependencies. To do so, LSTMs have *information gates*, allowing nodes to retain memory from relevant data while also forgetting unnecessary information to the final prediction (29). The architecture of a standard LSTM network is slightly more complex than the RNN, as it consists of a chain of repeating memory blocks (or nodes), each consisting of four layers. In brief, the forget, input and output gates update and control the memory cell via hyperbolic tangent and sigmoid activation functions. The information encoded in the new cell states allow the LSTM to capture long term dependencies and relations in sequential data, possibly rendering it the most effective DL algorithm for time-series analysis. Due to the aforementioned advantages of LSTMs over other algorithms, and the encouraging results of prior research in this field of study, this line of work implements such a model for the accurate classification and prediction of IOH events five minutes in advance during colorectal surgery.

To date, many algorithms depend on invasive arterial line wave-forms recorded using arterial catheters and pressure transducers, both of which are selectively performed on high-risk patients, therefore limiting the applicability of their approach. Most importantly, most models developed thus far do not consider the heterogeneity of the patient cohort used to train the algorithms. The need to develop IOH predictive models tailored to specific patient phenotype is crucial, as it would significantly enhance their predictive performance and ultimately prevent the clinical complications of (and not limited to) colorectal surgery. Thus a combination of deep learning and unsupervised learning (clustering) was considered. In this context, clustering can be used to stratify the patients and allow for more tailored and refined modelling. The diverse clinical characteristics of each patient play an important role in defining their risk of suffering from hypotensive events during a surgical operation. Due to this complexity, a simple and generic modelling technique might not be sufficient to capture heterogeneity; crucial for the accurate risk prediction of IOH. This study investigates the effect of training the LSTM model on different clusters of patients, defined by their similarity in parameters such as BMI, age, type of anaesthetic drug administered (e.g., ephedrine), preoperative hyponatremia and hypertension.

We furthermore investigate the possibility of clustering patients according to their time-series MBP data at the beginning of the operation, in an attempt to improve the predictive performance of the LSTM model. Distance measures used in standard clustering algorithms are not appropriate for time-series as they are invariant to time shifts and ignore temporal dependencies. For instance, Euclidean distance yields pessimistic similarity measures when it meets alterations in the time axis. In other words, it is not capable of identifying the similarity between two time-series if one of them is slightly shifted in time, even if they are highly correlated. Instead, Dynamic Time Warping (DTW) is a time-series similarity measure that finds the optimal non-linear alignment between two time-series irrespective of their time, speed and length, and thus is deemed more suitable for our analyses. For instance, given two time-series X and Y, DTW creates a wrapped path between each value in X and the closest point in Y, resulting in a sounder similarity assessment that minimises the Euclidean distance between aligned series (37).

## Results

Following data pre-processing, 604 out of 1106 colorectal surgery patients satisfied the inclusion criteria and were therefore considered for the analysis, of which 378 cases (62%) experienced at least one event of IOH. Data characteristics of the complete dataset, as well as within those having suffered, or not, from hypotension are detailed in Table 1. Due to the time-series nature of the dataset, the table indicates the number of observations belonging to each category, rather than the number of patients. The mean age of the cohort was 62, with the hypotensive group being slightly older than the average. Additionally, these patients were characterised by significantly lower BMI and weight, known to be inversely associated with the risk of hypotension. As expected, the non-hypotensive group had a 12% increased MBP compared to the hypotensive group.

**Table 1:** Population characteristics grouped by patients who experienced and did not experience IOH.

| Grouped by Status (hypotensive/normal) |  |  |  |  |  |
| --- | --- | --- | --- | --- | --- |
|  |  | Overall | IOH | Normal | P-Value |
| Clinical data, mean (SD) |  |  |  |  |  |
| N of observations |  | 2370396 (=604 cases) | 1733617 (=378 cases) | 636779 (=226 cases) |  |
| Age (years) |  | 62.4 (12.0) | 63.1 (12.3) | 60.5 (11.0) | <0.001 |
| Height (cm) |  | 162.0 (8.5) | 161.3 (8.5) | 164.1 (8.3) | <0.001 |
| Weight (kg) |  | 60.5 (10.7) | 59.5 (10.5) | 63.3 (10.8) | <0.001 |
| BMI (kg/m2) |  | 23.0 (3.4) | 22.8 (3.4) | 23.5 (3.5) | <0.001 |
| ASA physical status, n (%) | 1.0 | 641007 (27.0) | 445737 (25.7) | 195270 (30.7) |  |
|  | 2.0 | 1521098 (64.2) | 1160286 (66.9) | 360812 (56.7) |  |
|  | 3.0 | 170835 (7.2) | 108610 (6.3) | 62225 (9.8) |  |
|  | 4.0 | 7051 (0.3) |  | 7051 (1.1) |  |
| Preoperative hypertension, n (%) | No | 1482079 (62.5) | 1078356 (64.4) | 403723 (62.2) | <0.001 |
|  | Yes | 888317 (37.5) | 655261 (36.6) | 233056 (37.8) |  |
| Diabetes, n (%) | No | 2041528 (86.1) | 1500333 (86.5) | 541195 (85.0) | <0.001 |
|  | Yes | 328868 (13.9) | 233284 (13.5) | 95584 (15.0) |  |
| Preoperative haemoglobin (g/dL) |  | 12.1 (2.8) | 11.9 (2.7) | 12.6 (2.9) | <0.001 |
| Preoperative platelet count (x1000/mcL) |  | 259.6 (94.2) | 260.6 (100.5) | 256.8 (74.3) | <0.001 |
| Preoperative PT* (%) |  | 99.0 (20.1) | 98.8 (20.4) | 99.6 (19.5) | <0.001 |
| Preoperative aPTT** (sec) |  | 31.7 (6.7) | 31.6 (6.4) | 31.8 (7.3) | <0.001 |
| Preoperative Na (mmol/L) |  | 132.7 (32.7) | 133.1 (32.0) | 131.7 (34.6) | <0.001 |
| Preoperative K (mmol/L) |  | 3.8 (1.2) | 3.8 (1.2) | 3.8 (1.3) | <0.001 |
| Preoperative Glucose (mg/dL) |  | 114.9 (47.5) | 116.1 (49.8) | 111.7 (40.3) | <0.001 |
| Preoperative Albumin (g/dL) |  | 3.9 (0.8) | 3.8 (0.8) | 3.9 (0.9) | <0.001 |
| Preoperative Ast*** (IU/L) |  | 22.0 (13.8) | 22.2 (15.0) | 21.4 (9.7) | <0.001 |
| Preoperative Alt**** (IU/L) |  | 19.0 (15.0) | 18.6 (15.3) | 20.0 (14.3) | <0.001 |
| Preoperative blood urea nitrogen |  |  |  |  |  |
|  |  | 14.1 (6.4) | 14.2 (6.6) | 13.8 (6.0) | <0.001 |
| (mg/dL) |  |  |  |  |  |
| Preoperative Creatinine (mg/dL) |  | 0.8 (0.8) | 0.8 (0.7) | 0.9 (0.9) | <0.001 |
| Estimated blood loss (mL) |  | 220.2 (484.2) | 246.1 (552.3) | 149.7 (188.6) | <0.001 |
| intra-operative urine output (mL) |  | 188.3 (215.0) | 190.1 (225.4) | 183.6 (183.7) | <0.001 |
| intra-operative Crystalloid (mL) |  | 1001.3 (935.3) | 1036.3 (1022.6) | 905.9 (630.0) | <0.001 |
| intra-operative Propofol (mg) |  | 47.9 (55.8) | 44.6 (54.7) | 56.6 (57.7) | <0.001 |
| intra-operative Fentanyl, (mcg) | 0 | 1717352 (72.5) | 1285691 (74.2) | 431661 (67.8) | <0.001 |
|  | 100 | 546053 (23.0) | 362069 (20.9) | 183984 (28.9) |  |
|  | 150 | 11212 (0.5) | 6912 (0.4) | 4300 (0.7) |  |
|  | 200 | 27186 (1.1) | 19348 (1.1) | 7838 (1.2) |  |
|  | 50 | 68593 (2.9) | 59597 (3.4) | 8996 (1.4) |  |
| intra-operative Rocuronium (mg) |  | 87.0 (23.9) | 87.8 (24.8) | 84.9 (21.1) | <0.001 |

|  | Grouped by Status (hypotensive/normal) |  |  |  |
| --- | --- | --- | --- | --- |
| intra-operative Ephedrine (mg) | 10.5 (12.8) | 13.1 (13.5) | 3.5 (6.8) | <0.001 |
| intra-operative vital signs with Primus, mean (SD) |  |  |  |  |
| MAC***** (unitless) | 0.4 (0.5) | 0.4 (0.5) | 0.5 (0.6) | <0.001 |
| Positive End Exp. Pressure (mbar) | 1.8 (2.4) | 1.7 (2.4) | 2.2 (2.5) | <0.001 |
| intra-operative vital signs with Solar8000, mean (SD) |  |  |  |  |
| Body temperature (°C) | 33.8 (5.6) | 34.0 (5.3) | 33.3 (6.2) | <0.001 |
| ETCO2 (mmHg) | 33.4 (7.5) | 33.6 (7.1) | 32.8 (8.5) | <0.001 |
| FEO2 (%) | 38.9 (17.1) | 38.4 (16.5) | 40.4 (18.6) | <0.001 |
| FIO2 (%) | 44.3 (17.4) | 43.8 (16.8) | 45.7 (19.0) | <0.001 |
| HR (/min) | 71.2 (15.4) | 71.6 (15.6) | 70.2 (14.7) | <0.001 |
| INCO2 (mmHg) | 1.1 (1.5) | 1.1 (1.5) | 1.0 (1.5) | <0.001 |
| Diastolic BP (mmHg) | 69.8 (13.1) | 67.5 (12.6) | 75.9 (12.5) | <0.001 |
| Mean BP (mmHg) | 86.0 (16.9) | 83.2 (16.2) | 93.6 (16.2) | <0.001 |
| Systolic BP (mmHg) | 115.6 (23.2) | 112.1 (22.3) | 125.0 (22.9) | <0.001 |
| Plethysmography recorded HR (/min) | 71.6 (15.3) | 71.9 (15.3) | 70.9 (15.4) | <0.001 |
| Plethysmography recorded SPO2 (%) | 99.0 (1.9) | 99.0 (2.0) | 99.1 (1.8) | <0.001 |
| Capnography recorded Resp. rate (/min) | 13.8 (3.4) | 13.8 (3.3) | 13.7 (3.7) | <0.001 |
| Ventilator recorded Mean Airway Pressure (mbar) | 5.6 (2.8) | 5.4 (2.7) | 5.8 (3.0) | <0.001 |
| Ventilator recorded Resp. rate (/min) | 13.8 (4.0) | 13.9 (3.9) | 13.7 (4.3) | <0.001 |

### LSTM model

The univariate LSTM classification model was trained on 484 patients for the prediction of hypotensive events 5 minutes in advance, using 5 minutes of MBP data. Evaluation of the model performance on the test dataset yielded an Area under the ROC Curve (AUC) score of 0.831 (Figure 1) and AUPRC= 0.225 (Figure 2). The model calculates a vector containing a probability for every output segment *Y* of the test dataset. In other words, it predicts a risk score (*∈* [0, 1]) of a particular output segment *Y* being hypotensive. The closer the score is to 1, the higher the risk of that 5-minute-long segment suffering an IOH event (MBP *<*65mmHg).

**Figure 1:**
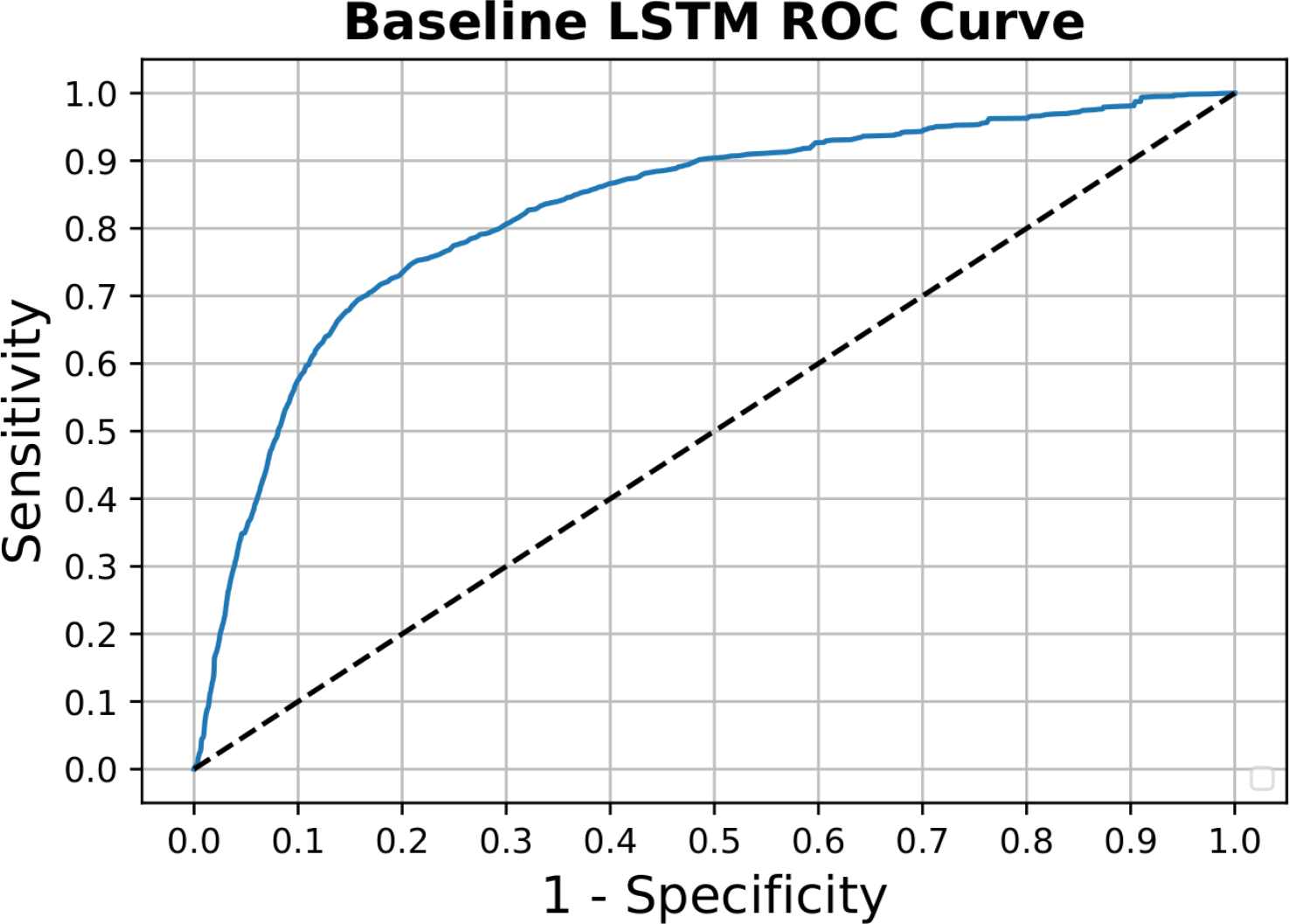
AUC-ROC plot for baseline model

**Figure 2:**
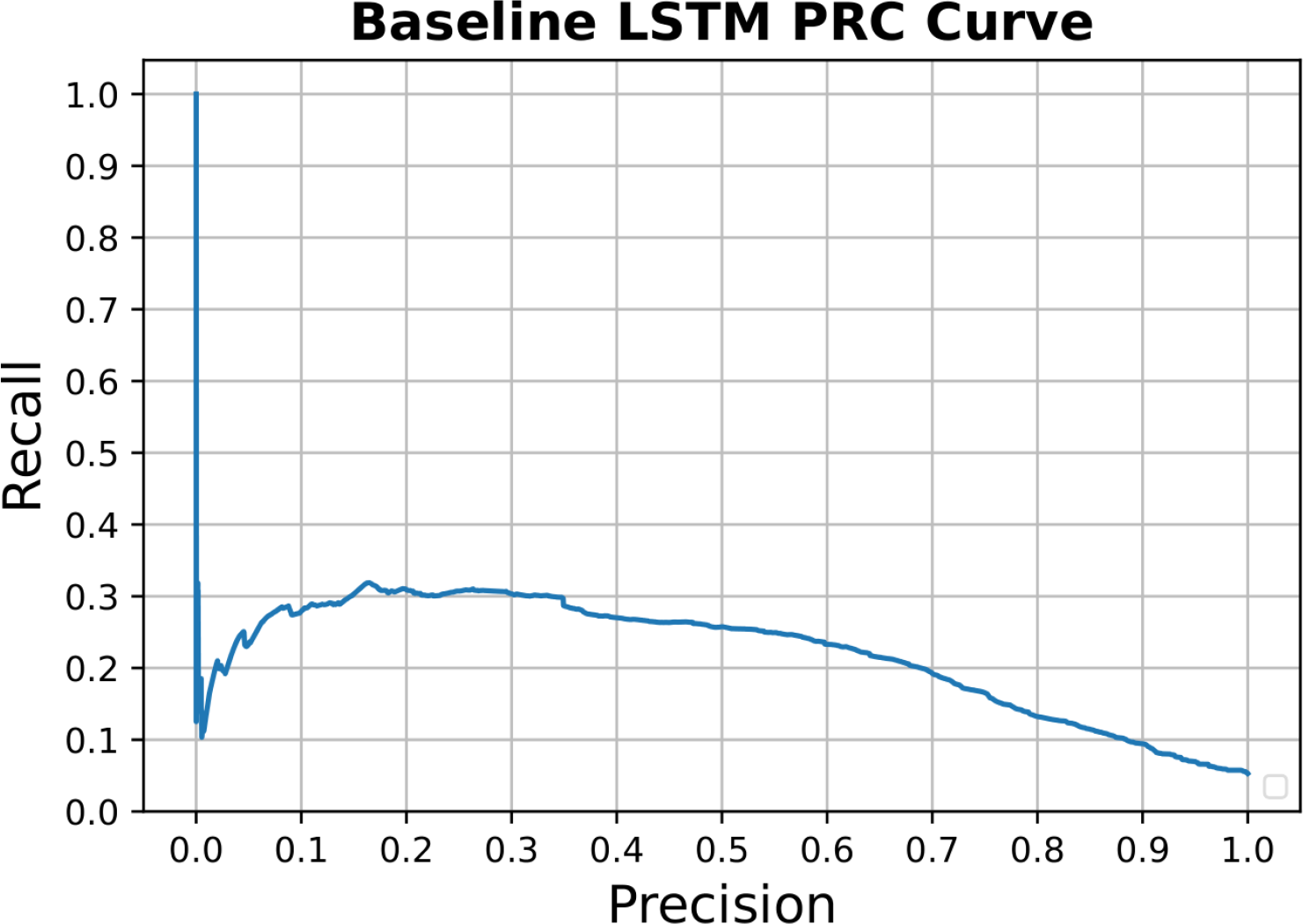
AUC-PRC plot for baseline model

Some examples of cases from the test dataset were visualised (Figure 3). The *x* axis represents the MBP of the patient, while the *y* axis represents the consecutive output segments *Y* generated by the time window method throughout the operation time. The vertical line at 65 mmHg denotes the threshold for IOH classification. The red line shows the true MBP of the patient at every output segment *Y*. The blue line can be interpreted as a warning score for the occurrence of hypotension at every output segment *Y*. As seen on the *x* axis on the right side of the plot, it is a probability value. The higher the blue line, the bigger the risk of that segment being hypotensive. When both lines cross the 65mmHg bar threshold simultaneously, that area of the graph becomes shaded, indicating that those segments have correctly been predicted as hypotensive by the LSTM model.

**Figure 3:**
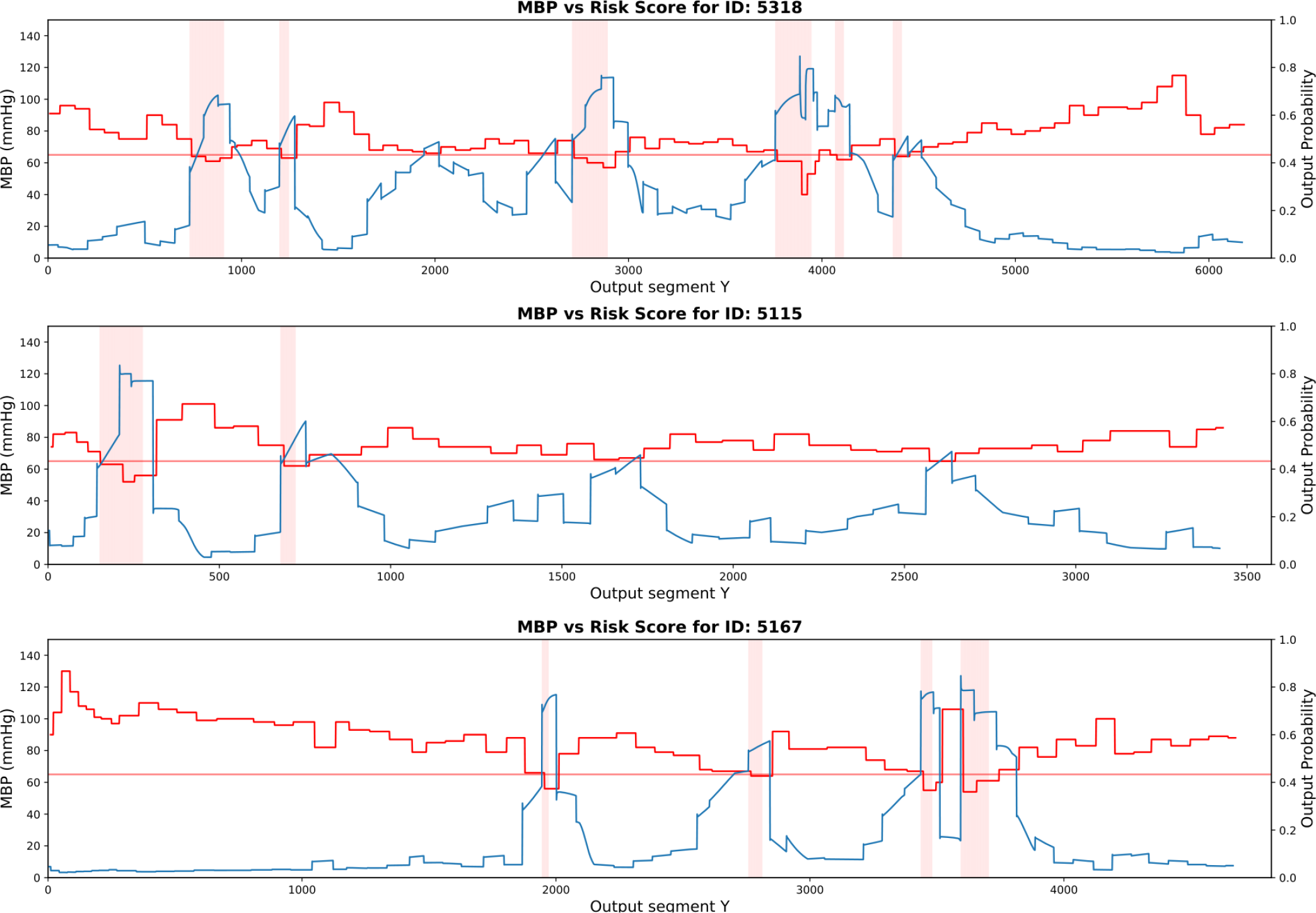
MBP vs Risk score plots for patients with case IDs: 5115, 5167 and 5318

### Clustering

The optimal number of clusters to group 604 patients based on their pre-clinical information is 3 based on the *Elbow* index (Figure 4). Performing K-means clustering (K=3) provides the following partition: cluster 0 composed of 36 patients (5.96%), cluster 1 composed of 348 patients (57,6%), and cluster 2 composed of 220 patients (36,4%) (Table 2). In addition, a polar plot was used to visualise the separation between clusters across clinical features (Figure 7). As expected, the main feature driving the differentiation was *preop_htn*, a categorical variable defining whether a patient had hypertension prior to the surgical operation. As seen from Table 2, 100% of cases in cluster 2 had preoperative hypertension, whereas 100% of cases in cluster 1 did not. Nearly one-third (1*/*3) of cases in cluster 0 suffered preoperative hypertension (30%).

**Table 2:**
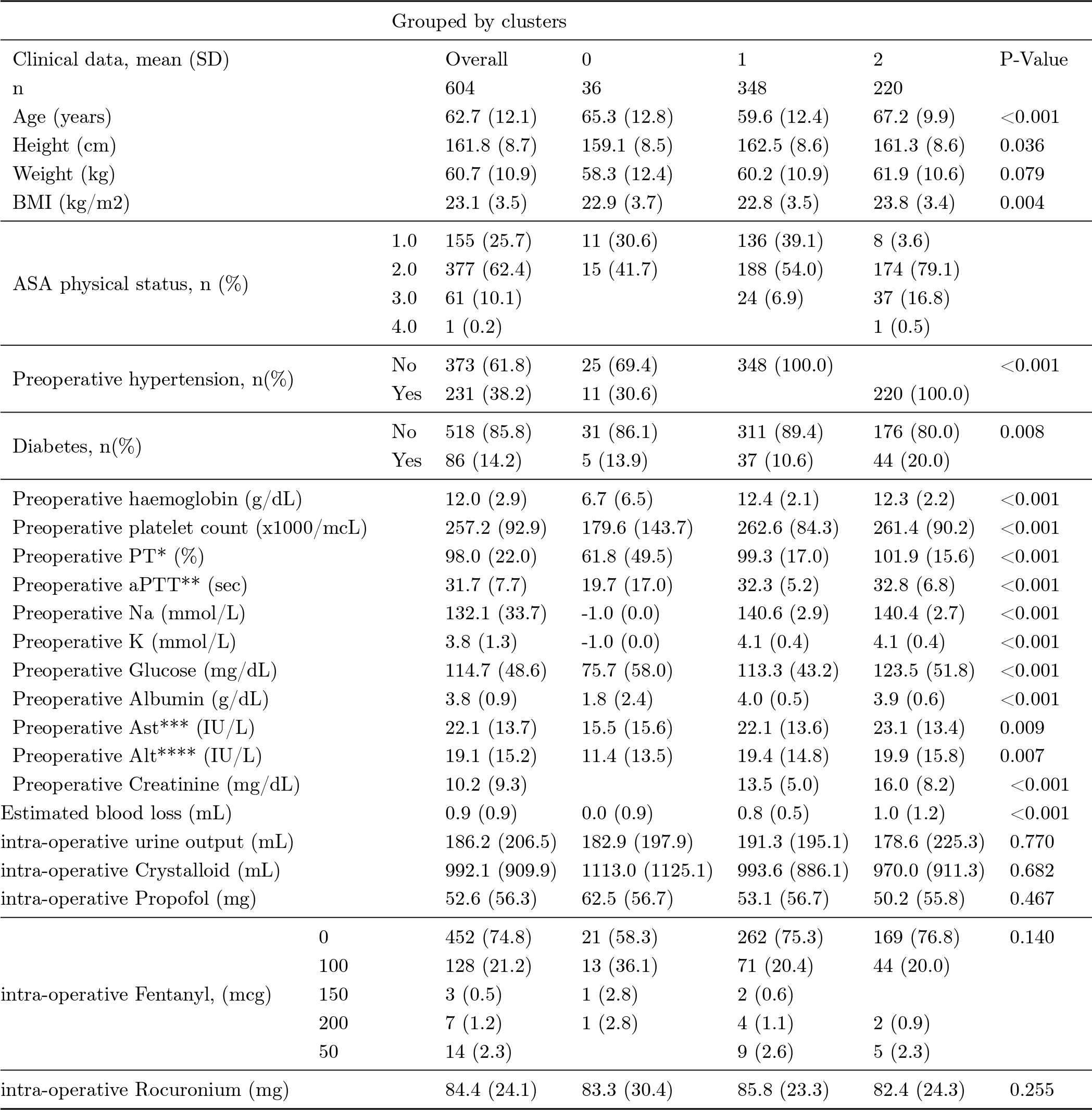
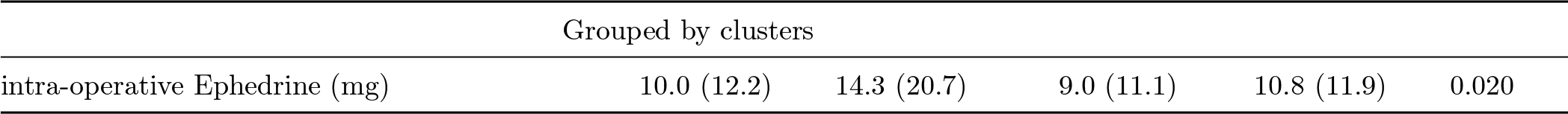
Preoperative population characteristics grouped by clusters.

**Figure 4:**
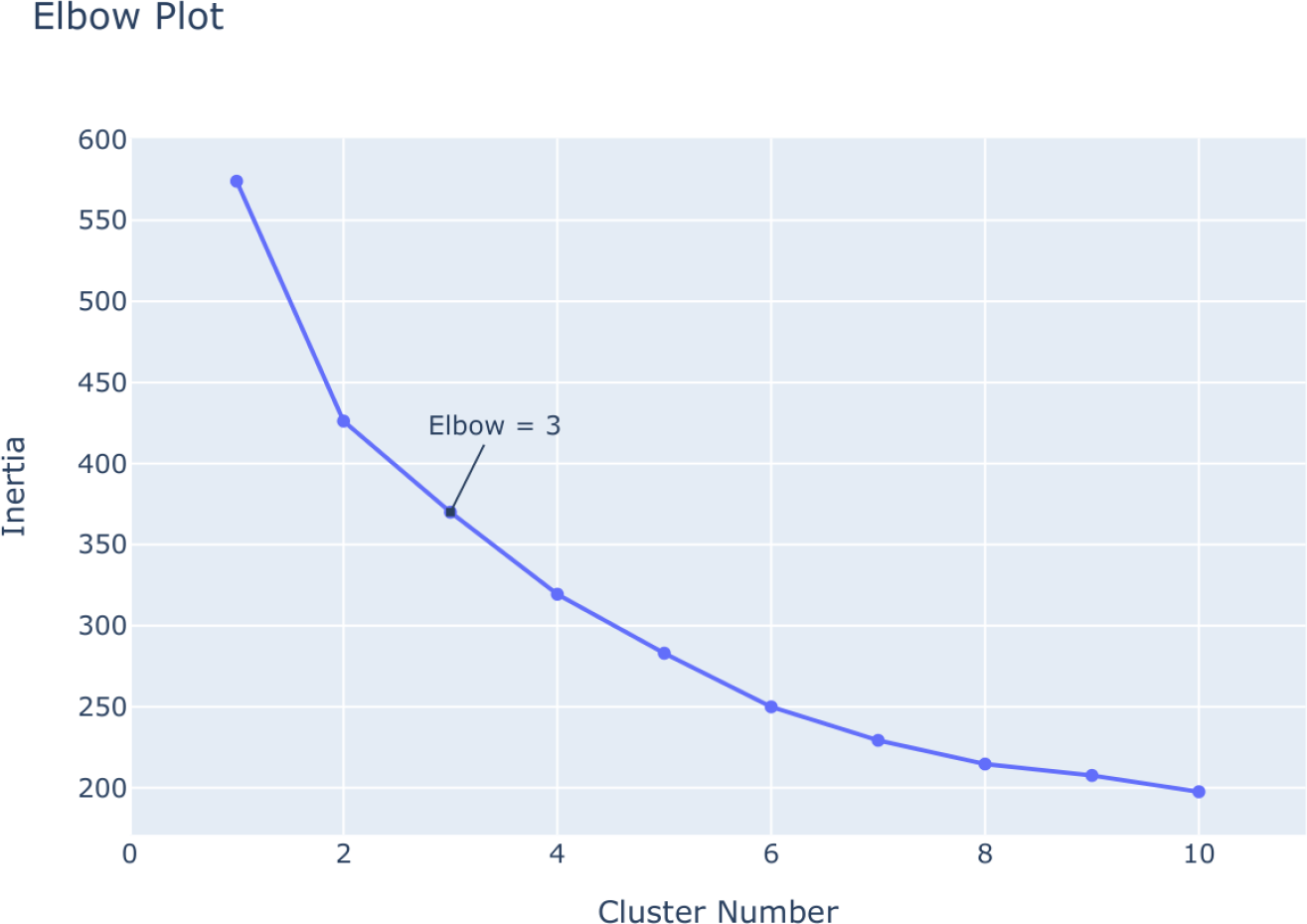
Elbow plot for K-means clustering

Another important driving factor was the *ASA physical status score*, with 37 out of 61 cases of class III belonging to cluster 2. Age also varied significantly among clusters, with a mean age of 59 years old being recorded for cluster 1, compared to 65 and 67 for clusters 0 and 2 respectively. Based on the polar plot, it can be observed that cluster 0 has grouped the few cases that are characterised by lower preoperative levels of albumin, haemoglobin and a lower platelet count, in addition to the aforementioned differences in terms of ASA status score, age and BMI. Training the LSTM model on each cluster gave the following results: i) cluster 0 yielded an *AUC* = 0.81 (*AUPRC* = 0.131), ii) cluster 1 yielded an *AUC* = 0.85 (*AUPRC* = 0.208), and iii) cluster 2 yielded an *AUC* = 0.822 (*AUPRC* = 0.254). Hence, when LSTM is applied to a specific cluster of patients characterised by the absence of pre-operative hypertension (i.e., cluster 1), the AUC is increased versus the baseline model (that is applied to the whole cohort without partitions).

### Sensitivity Analysis

As part of a sensitivity analysis, the first 100 recordings of MBP were used to cluster 604 patients. The elbow plot resulted in K=4 as the optimal number of clusters to differentiate patients accurately. This result was also supported by performing *Silhouette* analysis for K=4, which yielded the highest score (=0.30). As seen in Figure 5, the plots are more or less of similar thickness (except for cluster 2 which is of slightly smaller size) and show above average *Silhouette* scores. The labelled scatter plot on the right verifies these findings and allows for a better visualisation of the clusters. Applying the K-means algorithm (K=4) to MBP time-series with Euclidean distance yielded a *Silhouette* score of 0.30, whereas using Dynamic Time Wrapping (DTW) resulted in a slightly higher DBA *Silhouette* score of 0.33. We also see in Figure 6 the time-series trends that characterise each cluster post-normalisation. The four clusters were composed of 169, 157, 92 and 169 patients respectively. Again, the LSTM model was separately trained on each of these clusters, with the resulting AUC and AUPRC being compared to the baseline model in Table 3. Cluster 2, characterised by sudden drop in the MBP during the last moments of the predefined ∼ 3 minute period, yielded an AUC of 0.871 and AUPRC of 0.275. This represents a substantial increase versus the predictive performance of the baseline model (which yielded AUC=0.831 and AUPRC=0.225).

**Table 3:**
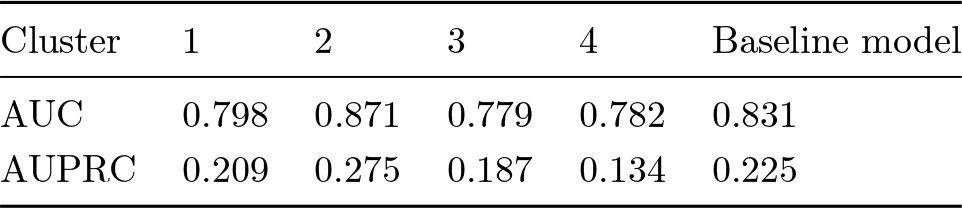
Results from LSTM with Sensitivity analysis.

**Figure 5:**
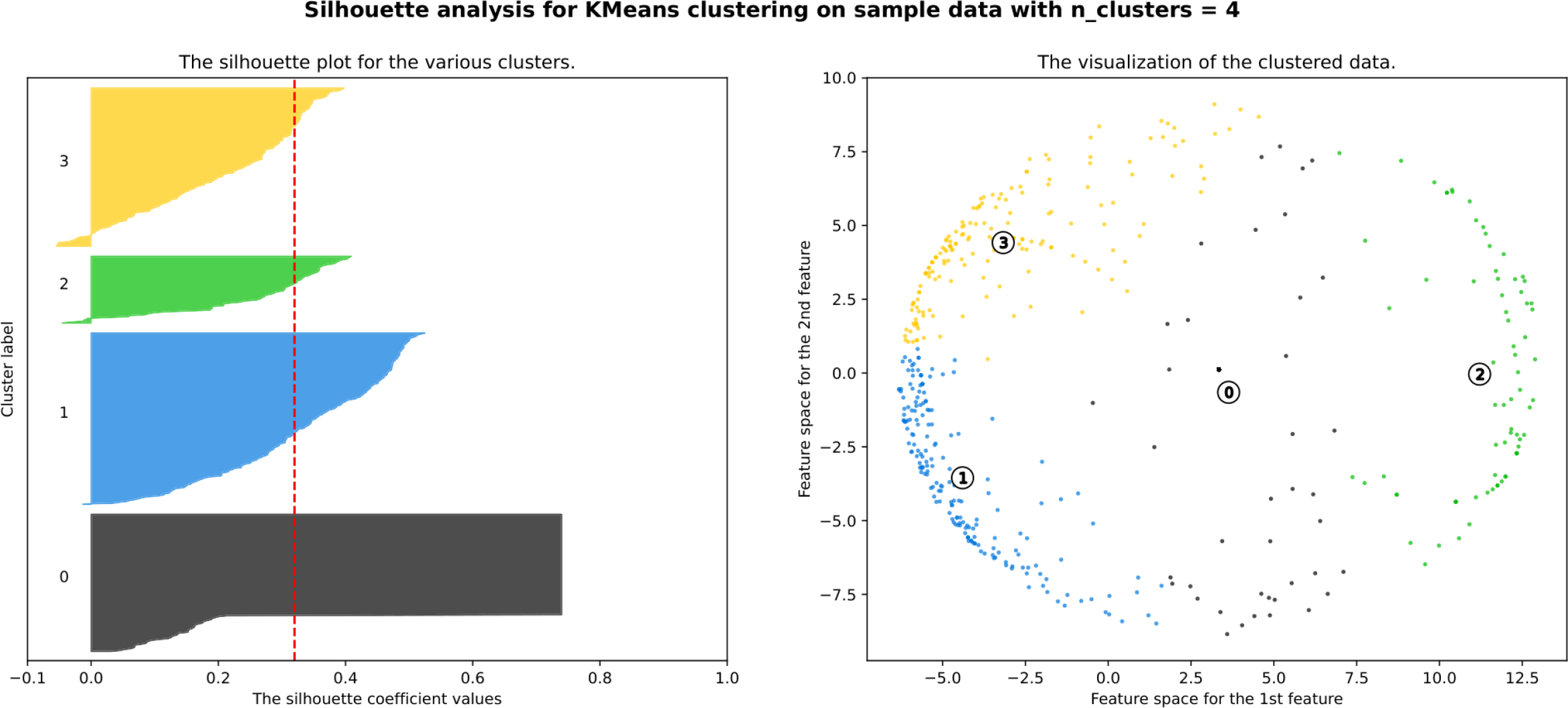
*Silhouette* plot (left) and scatter plot (right) for number of clusters=4

**Figure 6:**
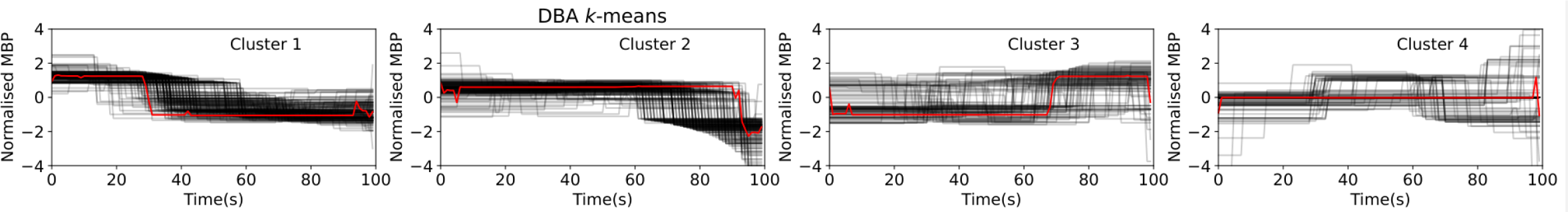
Plots of 100 first MBP values for each cluster in sensitivity analysis

**Figure 7:**
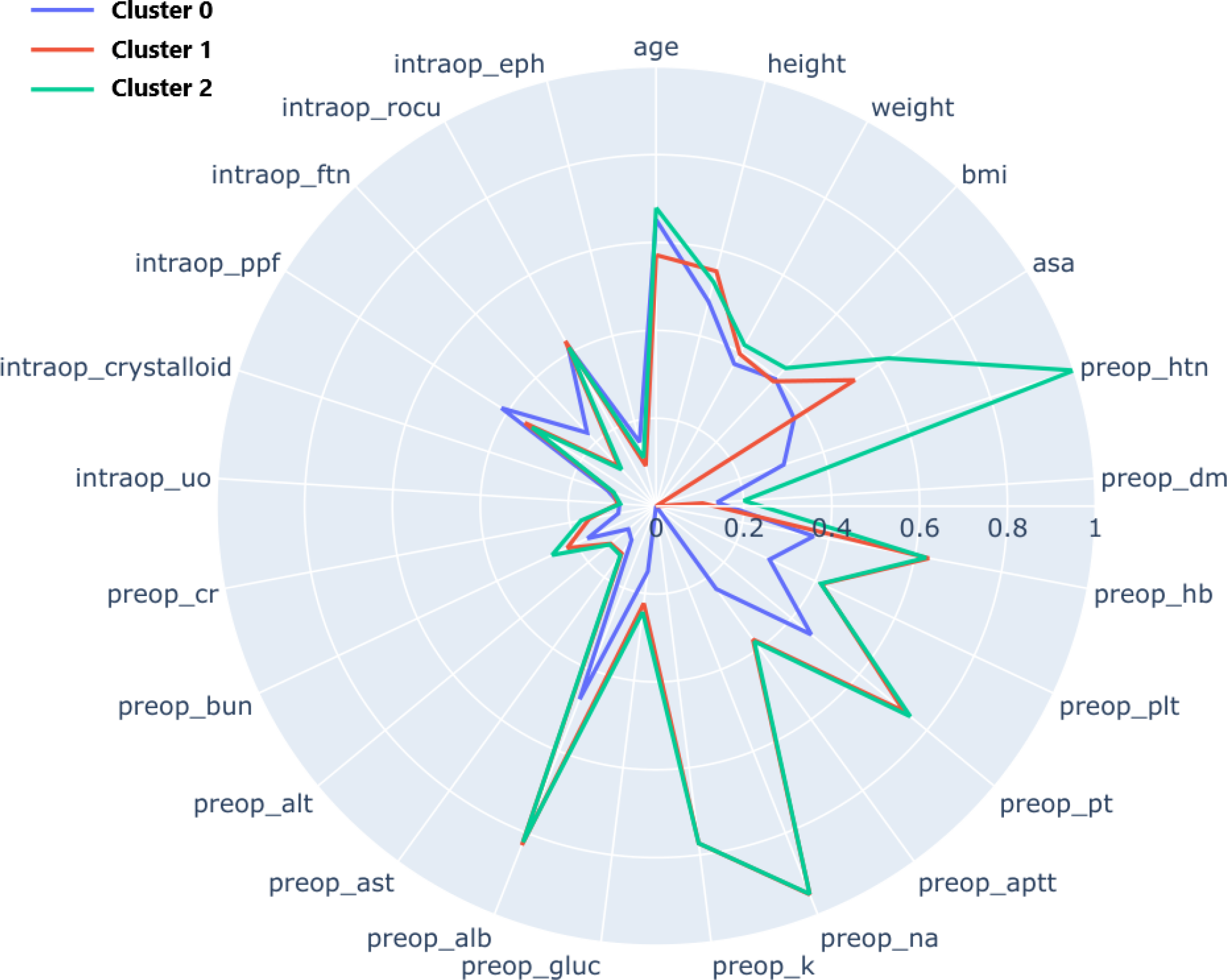
Polar plot for three clusters. Features are displaying peripherally and differences across clusters on the selected features using a scale from 0 to 1.

## Discussion

General anesthesia of patients undergoing surgery is often accompanied by the serious haemodynamic abnormality that is IOH. Currently, no reliable methods exist in clinics to predict that a patient will suffer a hypotension during an operation, although several devices are used to continuously monitor the patient’s vital signs in real-time. The existing need to minimise the post-operative complications of such events is what motivated this line of work in predicting the likelihood of a IOH events in patients undergoing colorectal surgeries. Importantly, our analysis tried to set up more realistic conditions than previous and related work, to increase the applicability and generalisation of the model to different settings and patients and to allow for more tailored model training using unsupervised learning. For instance, we used intra-operative vital signs collected using common monitoring devices present in most operating rooms, as well as MBP time-series data recorded in a non-invasive manner.

An LSTM classification algorithm was trained on samples of MBP generated via the sliding window method. Using 5 minutes of MBP time-series data, the algorithm was capable of classifying an IOH event five minutes in advance (AUC=0.831). Although the algorithm was trained offline on pre-recorded timeseries data, it could be used as a real-time prediction model for IOH in the operating room. By using the first five minutes of MBP data as input, the model would already be able to output the first IOH risk prediction score five minutes ahead. Running the model in an iterative fashion on input data using a one second step size would therefore yield a IOH risk score throughout the duration of the operation, similarly to how the model was tested on unseen patients. This risk score can greatly benefit anaesthesiologists who could be alerted of the proclivity towards an impending IOH event. As a result, clinicians could be given a substantial advantage in proactively treating the patient and averting the hypotensive event and the subsequent potential complications and Intensive Care Unit (ICU) stay.

Despite the encouraging results of this baseline predictive model, it is essential to better understand the different clinical phenotype of patients undergoing colorectal surgery to assess their risk of IOH. With the widespread adoption of EHR, much of this clinical data is already being collected and stored, but due to the expertise required to analyse it, it often remains underutilized. However, unsupervised machine learning offers the possibility to automatically identify underlying physiological patterns among a heterogeneous patient cohort. More precisely, unsupervised clustering techniques such as K-means can be used to automate the process of clustering patients according to their clinical characteristics, and evidently improve numerous ML models in predictive accuracy.

Thus, we trained the model on three clusters of patients that were different in the presence/absence of signs of preoperative hypertension, BMI, age, ASA physical status score, preoperative levels of haemoglobin, albumin and platelet count. Surprisingly, the type of anaesthetic agent used (e.g. propofol, rocuronium, ephedrine, fentanyl) did not influence patient clustering, which would have been interesting to investigate as these are known to influence IOH at different levels. All variables were selected via a thorough feature selection process according to their relevance in predicting MBP. Training the model on a particular cluster that has half the number of patients of the original dataset increased the predictive performance of the model with an AUC=0.85 (2,4% increase). In fact, a recent study demonstrates that patients categorised as ASA physical status III and IV are more prone to suffering from haemodynamic abnormalities such as IOH, as these patients are more susceptible to the vasodilatory effects of anaesthetic agents (1). Additionally, platelet count and haemoglobin level are known to be positively associated with blood pressure, rendering these important determinants of IOH risk, as was also seen in our results (38). Lastly, patients showing signs of preoperative hypertension are obviously less likely to suffer from IOH, which explains why this is a major driving factor in patient grouping. The fact that clustering patients according to clinical characteristics showed increased predictive performance, even if this was only observed in a single cluster, is a promising finding. This shows that repeating the analysis on a significantly bigger cohort while introducing a larger number of features could help increase the *Silhouette* score i.e. better matching of patients to their own cluster yielding more similar/compact clusters. This process could offer a more tailored approach to new incoming patients, as they would get categorised using similarity metrics with previous cases. Thus, the appropriate pre-trained model could be used on each incoming patient instead of applying a generic model that ignores clinical phenotype heterogeneity. Ultimately, recognising patterns in the clinical data of patients could potentially lead to better IOH prediction and therefore better post-operative outcomes.

Another unexplored yet exciting approach consists in the grouping of colorectal-surgery patients based on the patterns observed in their MBP some minutes after being put under anesthesia. Interestingly, performing this type of clustering did again increase the predictive performance of the baseline LSTM model by as much as 4.8% (AUC=0.871). To do so, the K-means algorithm was used with DTW, which is deemed as more appropriate for time-series classification than the standard Euclidean distance. The *Silhouette* score was increased, although not substantially, potentially explained by the fact that the time-series were of the same length (i.e., 100 values long) due to the nature of the problem. Nevertheless, this finding opens new avenues for future research. If time-series clustering were to be performed on a bigger cohort, the resulting clusters would be even more dissimilar. During the start of the operation, the trends characterising the MBP of a new incoming patient would be indicative of the appropriate pre-trained LSTM model to use in order to predict IOH during the rest of the operation.

This study contributes to the endeavor of limiting post-operative complications by demonstrating that routinely collected EHR and easily accessible vital sign data can be used to produce a IOH risk score using the LSTM classification algorithm. Most importantly, it showcases the until now unacknowledged importance of classifying patients according to their clinical characteristics as well as their trends in intra-operative time-series data, therefore opening new avenues of research the biomedical domain and precision medicine.

Furthermore, the association of IOH in combination with the presence of hypotension prior of operation and ICU admission has been demonstrated in Figure 8. Here, we calculated the percentage of time spent in hypotension during surgery and calculated the hazard probability against ICU stay stratified by the presence (or not) of the combinations of IOH and pre operation hypotension, using the relevant variables from the original dataset. More specifically, we engineered a new variable in the original dataset, that shows the percentage of the time during each patient’s operation where *MBP <* 65*mmHg*. The plot shows that there is a statistically significant association (*p* = 0.05) of hypotension pre and during the operation with ICU admission. Surprisingly, patients that exhibit no hypotension prior of surgery but IOH, are in bigger risk of admittance to ICU versus patients who had presence of hypotension prior of surgery too. These findings further support the significance of the capability to predict hypotension during surgery to better pro-actively avert ICU admission.

**Figure 8:**
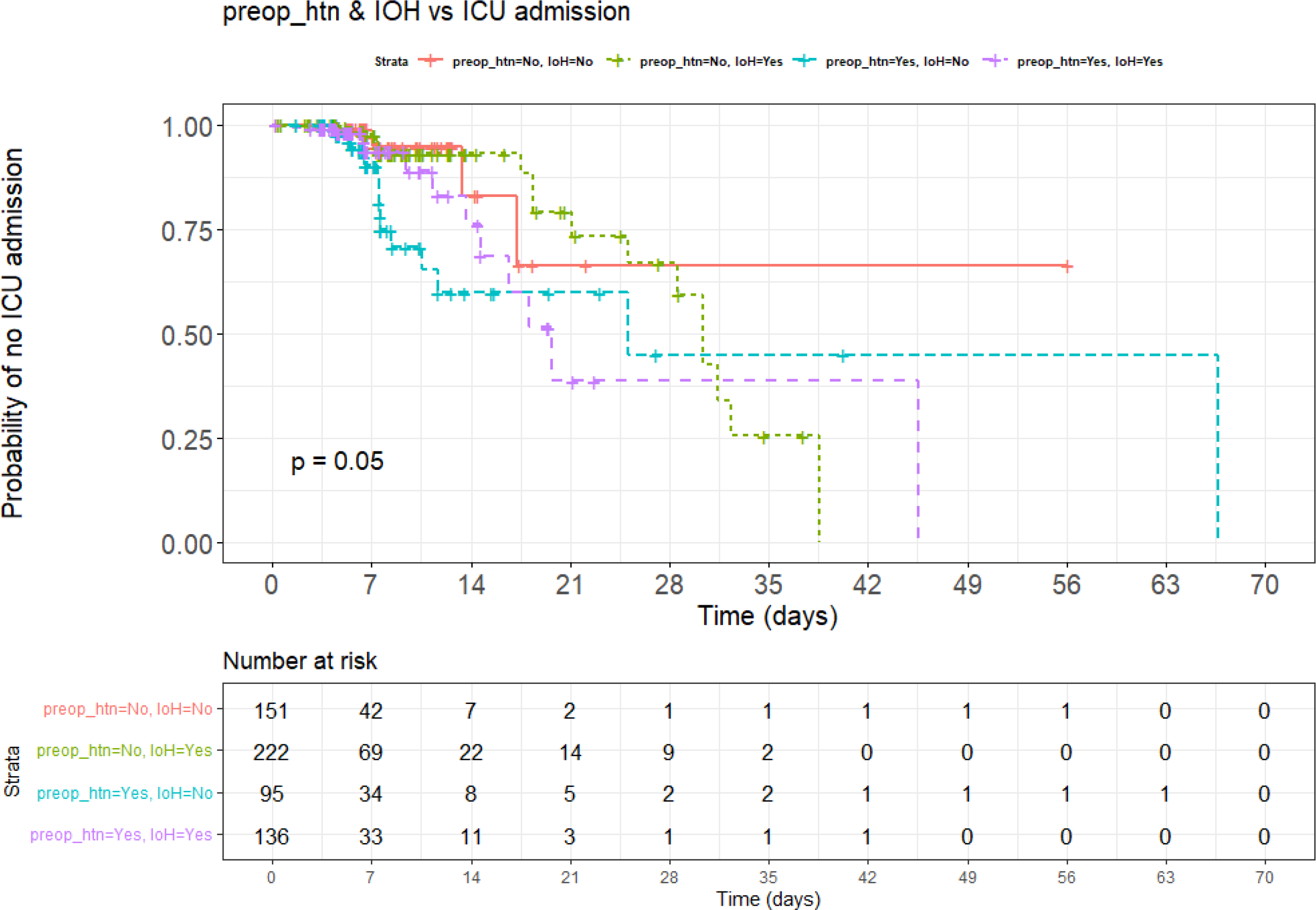
Association of ICU admission over IOH and pre-operation hypotension presence (discharge time in days). The legend includes all combinations between pre-operative hypotension (preop_ht) as present in the patient and intra-operative hypotension (IoH) each time. The p-value indicates the statistical significance of the score test of the null hypothesis, that there are no differences at all between the survival curves.

Although our approach is highly promising, it comes with some limitations. Our analyses were centred on colorectal surgery, with the patient cohort naturally consisting of similar cases. Therefore, our study cannot be directly applied to other types of surgeries, as these use different anesthesia procedures and drugs; however, a similar framework can be used if trained on the appropriate dataset and the models were fine tuned as required. The applicability of the model is also limited by the fact that it was trained and tested on a single cohort extracted only over a 14-month period and derived from a single institution and population. It would be interesting to perform training and external validation on institutionally independent patient cohorts, as well as cohorts from different populations, in order to increase model performance and generalisation.

This study also has potential for real-time prediction of hypotension, but the data streams were not actually analysed in real time. In fact, we do not cover exactly how anaesthesiologists would act upon the risk score predicted by the model. This implies the need for the development of a decision-support tool that could suggest interventions or treatment alternatives according to the risk score calculated by the model, tailored for the type of surgery and severity of the event of interest.

## Materials & Methods

We used publicly available data from *VitalDB* (30) (Vital Signs DataBase), published by Seoul National University Hospital and collected via the *Vital Recorder* (39), a software for the automated recording of time-synchronised data. VitalDB includes high-resolution multi-parameter data of 6388 non-cardiac surgeries taking place between June 2016 and August 2017. The dataset integrates intra-operative vital sign recordings collected from several monitoring devices with preoperative EHR data of patients. Each operation type has unique features and complications correlated to the nature of it, and usually follows a common sequence from anesthesia induction to incision. Therefore, this study focuses on adult patients having undergone colorectal surgery under general anesthesia. These not only comprise a major part of the surgeries taking place globally each year, but also often report instances of IOH. Although using invasive BP measurements would result in higher prediction accuracy, since data is continuously recorded, this study focuses on predicting IOH based on signals acquired during routine non-invasive monitors to expand the applicability of the model. More precisely, only intra-operative data collected via the Primus (anesthesia machine) and the Solar8000 (patient monitor) were used in our analyses.

Although there is no universal definition for IOH, adverse effects usually start occurring below a MBP threshold of 65 mmHg. Therefore, we define hypotensive events as rhythm segments with an *MBP <* 65*mmHg* lasting for longer than 1 min, whereas non-hypotensive events are defined as those with an *MBP <* 65*mmHg* stable for longer than 1 min. Due to the nature of the analysis (probability estimate), it is essential to have two easily separable and mutually exclusive labels. The original dataset contains intra-operative vital sign data and perioperative clinical information of 1106 colorectal surgery patients. More specifically, it consists of 101 features including: 1) patient demographic data, such as age, gender, ASA status score, 2) preoperative medical comorbidities, such as cardiovascular and respiratory risks, 3) intra-operative medications, 4) intra-operative vital signs, such as arterial blood pressure, heart rate and oxygen saturation. Vital signs were recorded at different time intervals according to the monitoring device, with a frequency of 2 seconds for numeric data recorded with Solar8000, and a frequency of 7 seconds with Primus recorded vitals. In order to harmonize the vital signs sampled at different rates, the time interval was set at 2 seconds, which is the frequency at which non-invasive MBP, the main event of interest, was recorded. In case of a missing data-point due to time difference, it was replaced with the last encountered non-null value using backward filling.

The initial selection of 101 features was decided based on clinical judgement of factors that are most likely to induce IOH, although ML was subsequently used to ensure unbiased feature selection. Among these features, 13 were removed due to having *>*80% missing values. Out of the 1106 patients that underwent colorectal surgery, two were excluded for the same reason. Blood pressure measurements that were beyond the physiological range were excluded according to the following criteria: (1) MBP *<*30mmHg or *>*200 mmHg, (2) SBP *<*50mmHg or *>*250mmHg, (3) DBP *<*20 or *>*160mmHg. In rare cases, some negative values were observed due to an error in the measuring equipment and were subsequently excluded, such as 61 negative values recorded using the Primus Positive end expiratory pressure (PEEP). Under-aged patients, as well as those having undergone a surgery lasting less than 2 hours were removed to obtain a more homogeneous cohort of patients with which to train the model.

We used correlation plots along with principal component analysis to identify highly correlated features. We merged the preoperative and intra-operative time-series datasets based on the patient ID, the EHR data was replicated for each instance of the patients’ operation. Finally, we imputed the missing values of the vital signs via linear interpolation.

We performed feature selection to prevent over-fitting and increase model performance. More precisely, dimensionality reduction was decided upon the output of three feature selection strategies: mean decrease in Gini, Boruta, and lastly Recursive feature elimination (RFE) using a RF classifier. The Gini index is an ensemble learner based on randomised decision trees that measures the decrease in the impurity of the selected features as each tree branches. A higher decrease in mean Gini index is indicative of a variable that contributes more to the sorting and grouping of the target variable (MBP) to accurately classify IOH events. The Boruta algorithm is a wrapper method also trained on a random forest classifier. However, it does so on a shuffled copy of the original features, called shadow features, and applies a feature importance measure such as mean decrease in accuracy. The main idea of this approach is that instead of competing among themselves, the features are compared to a randomised version of them. All variables declared insignificant due to low feature importance were removed, and the model is repeated until all variables are classified based on the selection threshold (40). Lastly, Recursive Feature Elimination (RFE) is an automatic feature selection wrapper method that uses subsets of features to train the model and allows for the addition or removal of them. Using a prediction error, RFE selects a minimal set of variables needed for an accurate predictive model (4). Following the implementation of these feature selection strategies, a total of 36 variables that were deemed unimportant for IOH prediction were dropped from the dataset. Intra-operative anaesthetic medications were regarded as continuous variables indicating the dose administered, whereas the value was converted to 0 in the case of no data availability. All other variables were continuous except for the existence of preoperative diabetes, preoperative hypertension, and ASA physical status score, all of which were categorical.

These pre-processing steps culminated in a final dataset composed of 42 variables and 604 patients having undergone colorectal surgery, for each of which the operation is recorded at two second intervals. This yields a total of 2.370.396 observations, each including the preoperative data (25 variables) and the intra-operative vital signs (17 variables) of a single second during the operation of each patient. Further technical information on the implementation of the LSTM model and clustering is given in the Supplemental Information.

## Data repository

All generated models and data are available on Mendeley DOI: 10.17632/f8bd3djyrd.1.

## Data Availability

All data are publicly available online at : https://vitaldb.net/dataset/

All models and pre/post processing of data are available at DOI: 10.17632/f8bd3djyrd.1.

https://vitaldb.net/dataset/

## Acknowledgements

MC-H acknowledges support from the H2020-EXPANSE (Horizon 2020 grant No 874627) and H2020-LongITools (Horizon 2020 grant No 874739).

## Supplemental Information

### The LSTM Model

Time-series data are a sequence of values that have been ordered by a time index. However, this format is not suitable for supervised learning, where the training process is comprised of input and output patterns, allowing the algorithm to learn how to map sequences of past observations as input to output observations. We converted the time-series data into sequences of observations following a sliding window approach in order to be compatible with the supervised learning. This step is essential to divide the dataset into sequences of observations from which the algorithm can learn. In a single step prediction, the data is re-framed for the prediction of a single output value. Nevertheless, multistep ahead predictions are more commonly used and of greater relevance to this study. All models were developed using Python (v. 3.9.1). The implementation of LSTM was facilitated by the *Keras* library and *TensorFlow*. The study aimed at implementing an LSTM algorithm for the accurate classification of hypotensive events 5 minutes in advance, using 5 minutes of time-series data. Since the dataset was shrunk to only contain MBP values recorded every 2 seconds, 5 minutes of operation duration are equal to 150 values. The window method was used for each patient separately as can be seen in Figure **??**. Samples of 330 values were generated with a sliding step size of 1 second. The first 150 values of the sample, known as the input, are the 5-minute period during which the data is used for model training. Next, the delay period of 5 minutes is how far into the future hypotensive events can be predicted, and the label of 1 minute (30 values) is how long the hypotensive event can be recognised for. A validation step was included to eliminate the effect of sudden drops or increases in MBP that are not a result of the patient’s own physiological response, but rather caused by artifact or external events. More precisely, samples with an absolute increase or decrease of more than 50 mmHg were excluded from the analysis. According to the patient’s operation duration, the sliding window generated a varying number of samples. Each sample is composed of two segments. Segment *X* represents the 5-minute input that is used to train the model into classifying the 1-minute-long segment *Y* (i.e., label) 5 minutes into the future. The segment Y can be classified as a i) hypotensive event ii) non hypotensive event, according to whether all MBP values in the segment are *<*65mmHg or not.

Next, the segments belonging to 80% of patients (n=484) were used to train the model, whereas the other 20% (n=120) were used to test the model and obtain performance metrics. We divided the data in batches (64) to facilitate the LSTM training. Input was normalised to zero mean and unit variance using Batch Normalisation (41).

The LSTM model was fit with a tensor consisting of input segments *X* as well as a tensor containing their corresponding output segment *Y* in each sample. The number of LSTM nodes was set at 16 as this yielded the most accurate predictions. A single Dense fully connected layer was used in the final stages of the neural network using a Sigmoid activation function. In our case, observations with an output close to 1 are more likely to indicate a hypotensive event, meaning that the segment *Y* is predicted to have all values *<*65mmHg, whereas an output close to 0 is indicative of a non-hypotensive event. Binary cross-entropy was used to compute the loss function by comparing each of the predicted probabilities to the actual class output. Also, the class weight parameter of the model was used to increase the weighting applied to the class indicative of a hypotensive event and thus address the issue of class imbalance.

When training DL models we used the Adam optimiser to increase model accuracy and reduce loss. The validation split, determining what fraction of the training data is used for validation, was set at 0.1 to evaluate the loss following every epoch. The number of epochs was set at 100 with the early stopping function set to 2, meaning that model training stopped after 2 consecutive epochs (i.e., complete passes of the training dataset through the LSTM) where the model performance did not improve on the validation dataset. The model was evaluated on the test dataset consisting of time-series data of new unseen patients. In terms of performance metric measures, the AUROC (area under the receiver operator characteristic curve) was to evaluate our binary classifier. The curve plots the true positive rate (TPS) vs the false positive rate (FPR) across different decision thresholds, with the AUC denoting the probability that the classifier will rank a random positive example higher than a random negative example (i.e. indicates the classifier’s ability to distinguish between classes). The area under the precision recall-curve (AUPRC) is another important performance metric for binary classifiers, particularly in the case of imbalanced datasets like ours. The graph shows the precision versus recall across different decision thresholds. However, it is slightly more difficult to interpret since the baseline, instead of always being 0.5 as in AUC (random classifier), is defined as the fraction of positives (number of positive examples divided by the total number of examples). In this analysis, the class indicative of hypotension represents 6.3% of the total, yielding a baseline AUPRC of 0.063.

### Clustering

K-means clustering was used to cluster the patient cohort composed of 604 cases according to their clinical characteristics. Due to the variables containing numerical features measured in different units, these must be transformed to the same comparable scale, as they could otherwise impact the algorithm performance. In this case, MinMax normalisation was used to scale data to a range of [0-1]. Then, the optimal number of clusters was obtained via the Elbow index to fit the K-means algorithm on a total of 25 variables. We then applied the LSTM model to each of these clusters of patients separately with the same parameters.

To assess whether early patterns of MBP can be used to cluster patients in a more meaningful way for IOH prediction, K-means clustering algorithm was also applied to time-series data. The optimal number of clusters K was defined via both the Elbow plot and the *Silhouette* score. The algorithm was applied in a matrix containing the first 100 readings of the MBP for each patient. K-means was run twice, once using Euclidean distance and once using DTW (with DBA) in order to compare the *Silhouette* scores of both metric measures. Again, each cluster was used separately to train the LSTM algorithm at a later stage.

